# Bayesian Joint Spatiotemporal Modelling of Primary and Recurrent Infections of HFMD at County Level in Jiangsu, China, 2009–2023

**DOI:** 10.1101/2025.03.13.25323933

**Authors:** Weijia Wang, Hong Ji, Yifan Tang, Hongfei Zhu, Wendong Liu, Kai Wang, Liguo Zhu, Chengxiu Ling, Changjun Bao, Ying Wang

## Abstract

Over the past decade, multiple outbreaks of hand, foot, and mouth disease (HFMD) have occurred in East Asia, especially in China. It is crucial to understand the distribution pattern and risk factors of HFMD while also studying the corresponding characteristics of recurrent infections. This paper aims to jointly analyze the spatiotemporal distribution and influential factors of primary and recurrent HFMD in Jiangsu province, China, under the Bayesian framework. Using county-level monthly HFMD counts from 2009 to 2023, we proposed four spatiotemporal hierarchical models with latent effects shared in the reinfection sub-model to evaluate the influence of air pollution, meteorological factors, and demographic characteristics on HFMD on primary and recurrent HFMD infections. The integrated nested Laplace approximation (INLA) approach estimates model parameters and quantifies the spatial and temporal random effects. The optimal model with spatial, temporal, and spatiotemporal interaction effect indicates a significant positive influence of NO_2_, wind speed, relative humidity, and solar radiation, as well as a significant negative effect of PM_2.5_, O_3_, temperature above 27 °C, precipitation and COVID-19, on both infections. Scattered status and critical primary infection significantly positively affect both primary and recurrent incidence. Positive sharing coefficients reveal similar spatiotemporal patterns of primary and recurrent incidence. Non-linear analysis further demonstrates the influence of air pollution and meteorological factors. Our findings deepen the understanding of primary and recurrent HFMD infections and are expected to contribute to developing more effective disease control guidelines.

## 1 Introduction

Hand, Foot, and Mouth Disease (HFMD) is a contagious viral infection caused by enteroviruses, primarily affecting children under five years of age. Transmission occurs through contact with infected individuals or contaminated objects (U.S. Centers for Disease Control and Prevention, 2024). Symptoms typically appear after an incubation period of 3 to 7 days, including fever, oral ulcers, and rashes on hands and feet, with most patients recovering within a week (Guerra et al., 2017). While most cases are self-limited, severe infections can lead to neurological complications and even death (Legay et al., 2007). HFMD was classified as one of the notifiable infectious diseases after a large outbreak in 2008 originated in Fuyang City, Anhui Province (Wang et al., 2017; Zhang et al., 2010).

Numerous studies have identified various external factors influencing the incidence of HFMD, including meteorological conditions (Zhu et al., 2024), socioeconomic factors (Liu et al., 2024), and air pollution (Qian et al., 2023). For instance, higher temperatures generally increase the risk of HFMD, while air pressure negatively affects incidence (Xu et al., 2021). Socioeconomic variables, such as urbanization rate, gross domestic product (GDP), and population density, further shape disease dynamics by influencing population behavior and transmission patterns (Jiang et al., 2023). Air pollution has also been shown to play a critical role, with elevated NO2 levels linked to higher HFMD risk, while O3 exposure appears to reduce risk (Zhu et al., 2024). Interestingly, the relationships between these factors and HFMD incidence are often non-linear and context-dependent. For example, temperature impacts can vary by region, research by Bauer and Wakefield (2018) showed that temperature HFMD risk increases with temperature below 8.1 °C, but the effect becomes negative above this threshold. In contrast, in regions identified as cold spots, HFMD incidence remains stable or even decreases at temperatures exceeding 25 °C (Zhang et al., 2020). Similarly, air pollutants like PM_10_ exhibit non-linear effects too, with HFMD risk peaking at moderate concentrations before declining (Zhu et al., 2024).

Repeated infections with HFMD are common, disrupting patients’ daily activities and increasing the risk of further transmission, which has been a focus of epidemiological research (Zhong et al., 2022). Studies have explored reinfection risk factors, virus strains, and outbreak patterns. For instance, Huang et al. (2018b) analyzed the epidemiological and virological features of recurrent HFMD, applying the Kaplan-Meier method to estimate recurrence with different virus types and revealing risk factors associated with severe reinfection cases. Similarly, Peng et al. (2018) utilized the Cox proportional hazard model to exhibit the association between demographic characteristics, the type of virus in primary infection, and the risk of reinfection. Despite multiple research on HFMD primary and recurrent infection, there are few effort made to explore the spatiotemporal variability of recurrent HFMD, considering what Tian et al. (2018) did to study the overall incidence. Furthermore, although risk factors of primary and recurrent infections have been proposed in various studies, we still lack sufficient understanding of the relationship or comparison between the impacts on risk factors of two types of infections.

Many epidemiological studies have used spatiotemporal models under the Bayesian framework to capture disease patterns’ spatiotemporal variability (Zhu et al., 2023). This approach enhances statistical inference by incorporating prior knowledge with observed data. It allows for specifying spatial, temporal, and spatiotemporal random effects to study disease transmission and risk factors (Bolstad and Curran, 2016; Wang et al., 2024). The joint modeling approach effectively captures interdependencies and spatiotemporal dynamics by sharing latent effects between the two interest factors (Palmí-Perales et al., 2023). This framework has already been applied in environmental research (Pan et al., 2024), and we innovatively apply this Bayesian joint model to the study of primary and recurrent HFMD infections.

Using surveillance data from Jiangsu province, China, from 2009 to 2023, this study jointly analyzes primary infections and reinfections of HFMD, examining their spatiotemporal distribution patterns and risk factors, including meteorological variables, air pollution, and demographic characteristics. These variables are modeled as fixed effects in four joint Bayesian hierarchical models with varying spatial and temporal random effects. The joint modeling accounts for the nonlinear relationships of meteorological and air pollution factors. The optimal model is selected based on multiple Bayesian inference criteria, and comparing standard deviations of coefficients highlights the benefits of joint modeling by capturing the spatiotemporal dependence of HFMD incidence and exploring connections between primary and recurrent infections.

## 2 Materials

### 2.1 Study area and population

We obtained data on HFMD cases in Jiangsu from the National Infectious Diseases Reporting System (NIDRS), which has covered all the cases in Jiangsu province since 2009. Jiangsu is an eastern-central province of China (116°21′ − 121°56′E, 30°45′ − 35°08′N) and covers an area of 107.2 thousand square kilometers and 85.26 million residents by the end of 2023 (Jiangsu Provincial People’s Government, 2024). The climate in Jiangsu is predominantly humid subtropical, with a shift to a humid continental climate in the northern part of the province. Jiangsu is administratively divided into 13 cities, which include 95 county-level regions, with vector boundaries for the study area (Figure 1) obtained from the primary geographic database in the National Catalogue Service For Geographic Information of China.

**Figure 1:**
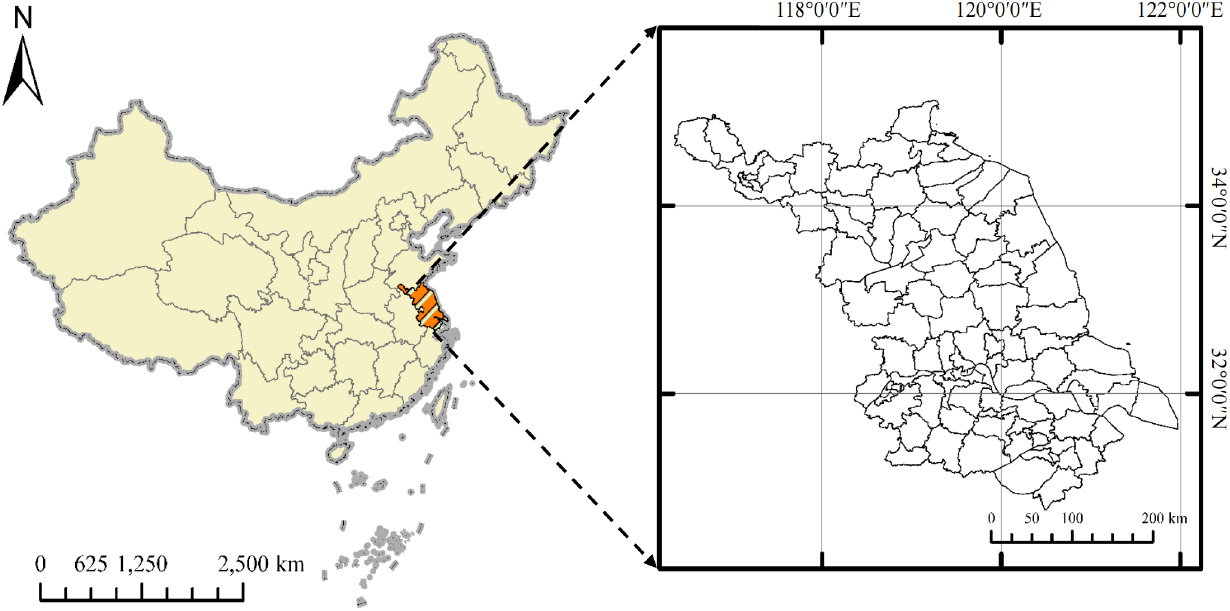
Geographical location and administrative division boundary map of Jiangsu province, China.

The original dataset contains a total of 1,578,702 infection cases reported from 2009 to 2023 in Jiangsu province, including patients’ names, ages, sex, dates of birth, type of accommodation, address type (local prefecture, other prefecture, other provinces), onset date, diagnostic date, an indicator for critically ill, etc. After excluding invalid recurrent cases (repeated primary infection, *n* = 7,615, 0.48%), cases of patients with addresses outside of Jiangsu Province (*n* = 44,653, 2.83%), and cases from patients with unspecified personal addresses (*n* = 786, 0.05%), we identified a total of 1,525,648 cases of HFMD who lived in Jiangsu Province from 2009 to 2023.

### 2.2 Outcomes

We aim to analyze primary and recurrent infections and categorize the data into primary and reinfection cases. Firstly, patients’ names are combined with dates of birth to create a unique ID for each individual. Then, cases that occur more than once are filtered as potential reinfection candidates. Finally, recurrent infections are identified if they occur at least 21 days after the last infection, which is considered valid according to previous studies (Hong et al., 2012; Huang et al., 2018b; Shi et al., 2018).

Among 1,525,648 cases, 1,477,294 were categorized as primary infections and 48,354 as valid reinfections. The address codes from the dataset were cross-checked with the county codes in the shapefile to ensure they match the current administrative divisions. Ultimately, we aggregated the data at the monthly and county levels, forming the spatial and temporal resolution for the study. Yearly population data in each county is obtained from the Statistics Yearbook released by each prefecture’s Bureau of Statistics and is recorded with a unit of 100,000.

### 2.3 Explanatory variables

Our analysis selects eleven risk factors as explanatory variables in the spatiotemporal model (Table 1), including five meteorological factors (Fu et al., 2019; Zhu et al., 2024), three air pollution sources (Qian et al., 2023), two demographic characteristics (Zhong et al., 2022), and one COVID-19 indicator (Jia et al., 2024). We standardized all numerical variables except temperature to ensure comparable fixed effects.

**Table 1:** Description for explanatory variables used in the spatiotemporal joint model.

| Type | Variables | Mean (Std) | (Min, Max) |
| --- | --- | --- | --- |
| Air pollution | PM<sub>2.5</sub> ($\mu\text{g}/\text{m}^3$) | 52.03 (22.98) | (12.63, 163.06) |
| | O<sub>3</sub> ($\mu\text{g}/\text{m}^3$) | 103.01 (31.00) | (33.84, 197.30) |
| | NO<sub>2</sub> ($\mu\text{g}/\text{m}^3$) | 34.79 (12.35) | (9.70, 112.07) |
| Meteorological factor | Temperature ($^{\circ}\text{C}$) | 16.97 (8.77) | (−4.73, 32.42) |
|  | Precipitation (mm) | 1.62 (1.53) | (0, 10.77) |
|  | Relative humidity (%) | 36.30 (8.06) | (36.30, 120.36) |
|  | Wind speed (m/s) | 1.23 (0.53) | (0.01, 4.15) |
| | Solar radiation ($\text{MJ}/\text{m}^2$) | 9.95 (2.48) | (4.02, 16.30) |
| Demographic characteristic | Critically ill (%) | 0.07 (0.25) | (0, 6.15) |
|  | Scattered status (%) | 6.64 (10.03) | (0, 210.81) |
| — | COVID-19 | — | (0, 1) |

Meteorological variables originate from ERA5-Land hourly data from 1950 to present (Sabater, 2019). We download hourly data daily between 2009 to 2023 and calculate the 24-hour average as a daily average, with the original spatial resolution of 9 km × 9 km. We select PM_2.5_ (Wei et al., 2020, 2021), O_3_ (Wei et al., 2022a) and NO_2_ (Wei et al., 2022b, 2023) as air pollution variables from the Global High Air Pollutants (CHAP) dataset, which provides monthly data at a 1 km × 1 km resolution. We averaged grid meteorological and air pollution data within each administrative boundary to obtain county-level data.

The two demographic explanatory variables include the proportion of critically ill individuals (patients diagnosed as critically ill for their first infection or reinfection in each county each month) (Shi et al., 2018) and scattered individuals (those not attending kindergarten or school in their county each month) (Zhong et al., 2022). Considering social isolation and potential under-reporting due to intense COVID-19 transmission from 2020 to 2022, we introduce an indicator variable for the COVID period (1 = COVID, 0 = non-COVID). The COVID period is the three years from January 2020, encompassing the early outbreak and lockdown phases, to December 2022, when the transition to reopening occurred, as outlined in relevant documents (Ge, 2023).

## 3 Methods

### 3.1 Spatiotemporal joint models

We establish a joint Bayesian spatiotemporal model to analyze primary and recurrent HFMD infections and their influencing factors, simultaneously fitting two sub-models, incorporating shared random effects, and capturing spatiotemporal dependencies between the two infection types.

We conducted a dispersion test for the generalized linear model on primary and recurrent infection count data, which revealed significant overdispersion in the response. Additionally, 49.6% of the reinfection data are zero, highlighting the necessity to properly deal with excess zeros in the model (Agarwal et al., 2002; Arab, 2015). Therefore, we use negative binomial (NB) regression for primary infection cases (*Y*_0_(*s, t*)) and zero-inflated negative binomial regression (ZINB) for the reinfection cases (*Y*_1_(*s, t*)) at location *s* in time *t*. Our primary focus is on how the average number of primary infections and the truncated reinfection cases (denoted *µ*_0_(*s, t*) and *µ*_1_(*s, t*)) vary dynamically across locations while keeping constant scale parameters (*ψ*_0_, *ψ*_1_) involved in the NB and truncated ZINB models:

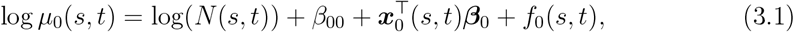

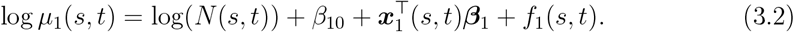

Here *N* (*s, t*) refers to the population at location *s* and time *t*, with its log transformation serving as the offset term. The intercepts are denoted by *β*_00_ and *β*_10_, while ***β***_0_ and ***β***_1_ represent the fixed effects associated with covariate sets ***x***_0_ and ***x***_1_ from Table 1. We also specify several random effects, *f*_0_ and *f*_1_, for comparison, as detailed in models 1 to 4. In these models, the spatial random effects are modeled using the Besag-York-Mollié 2 (BYM2) model (Simpson et al., 2017), and the temporal random effects are modeled using the first-order Gaussian random walks (RW1) model (Gómez-Rubio, 2020), with further details provided in Appendices A.1 and A.2, respectively.

#### Model 1: Mixed effects with only temporal random effect

We first consider a temporal random effect as the random effect in Eq.(3.1) as follows:

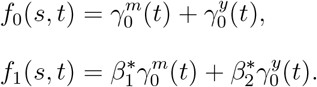

Here, 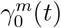 and 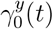 refer to the monthly and yearly latent effect at time *t*. The monthly random effect is defined to be cyclic, which illustrates the transition and similarity between December and January. The random effects are copied with flexibility that 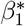 and 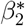 are the sharing coefficients linking the random effects in two sub-models. Meanwhile, 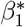 and 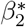 reflect the potential similarities or differences between the random variations in the two sub-models. For example, a significantly positive 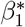 indicates that the monthly primary and recurrent infection variations follow similar patterns.

#### Model 2: Mixed effects with only spatial random effect

The second model includes fixed effects and a spatial random effect:

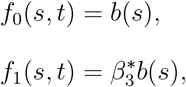

where *b*(*s*) refers to the spatial latent effect described by Eq.(A.1) that consists of a mixing parameter *ϕ* and a precision *τ*_*b*_ measuring the spatial variation proportion and total variation, and the sharing scale 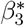 from NB to truncated ZINB model reflects the similarity or difference of the spatial effects between two submodels.

#### Model 3: Mixed effects with spatial and temporal random effects

Model 3 combines Models 1 and 2 above, taking both spatial and temporal random effects into account

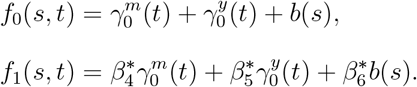

#### Model 4: Spatiotemporal interaction

Finally, we consider increasing Model 3 with an additional independent spatiotemporal interaction term representing different monthly variations at different locations

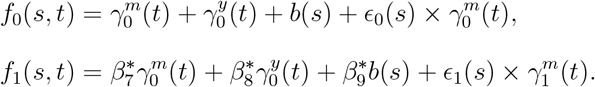

Here, we include two independent interactions of the unstructured spatial effect *ϵ*_*j*_(*s*) and the monthly random effect 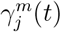 in the two submodels (i.e., *j* = 0, 1 for the NB and truncated ZINB models), by assuming that the monthly variation at each location is independent of other locations.

All statistical analyses were conducted using R software (version 4.4.1), with hyperparameters following the INLA default prior distributions.

### 3.2 Non-linear analysis

In our joint modeling framework, we consider the nonlinear effects of each air pollution and meteorological factor on primary and recurrent HFMD infections. Although frequentist spline methods and Bayesian latent Gaussian models differ fundamentally, with one relying on maximum likelihood estimation and the other on stochastic process modeling (Scheipl et al., 2013), a conceptual ‘equivalence’ exists between ordinary cubic splines and the second-order Gaussian random walks (RW2) process model. This equivalence is predicated on the scenario where an RW2 process is observed with additive independent Gaussian noise. In such cases, the posterior mode of the RW2 process aligns with the minimizer of a least squares problem that incorporates a cubic spline penalty.

### 3.3 Sensitivity analysis

We conduct two sensitivity tests to examine the robustness of our primary findings. First, for the spatial and temporal model parameters, we employ the penalized complexity (PC) priors (Simpson et al., 2017) that penalize the complex models to avoid overfitness. We set Prob(*τ*_RW_ *<* 1) = 0.01 for the RW1 model, which means the probability of the standard deviation 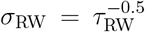 larger than 1 is very low. Second, for the BYM2 model, we use Prob(*τ*_BYM_ *<* 1.6) = 0.01 to penalize model with large variance and Prob(*ϕ >* 0.1) = 0.80.

## 4 Results

### 4.1 Descriptive analysis

Figure 2 (a) demonstrates the temporal variation of primary and recurrent incidence from 2009 to 2023. Since 2011, both infections have displayed similar seasonal patterns, peaking in April–June and October–December. Incidence levels generally increased from 2009 to 2018, declined from 2019 to 2022 during COVID-19, and rebounded in 2023.Figures 2 (b) and (c) display the spatial distribution of average incidence between 2009 and 2023 for primary and recurrent infection, respectively. In general, both types exhibit similar spatial patterns, with higher incidences in southern counties, where primary and recurrent incidences approach 20 and 1 per 100,000, respectively. In contrast, northern counties show lower incidences, with primary and recurrent incidences below 5 and 0.25 per 100,000, respectively.

**Figure 2:**
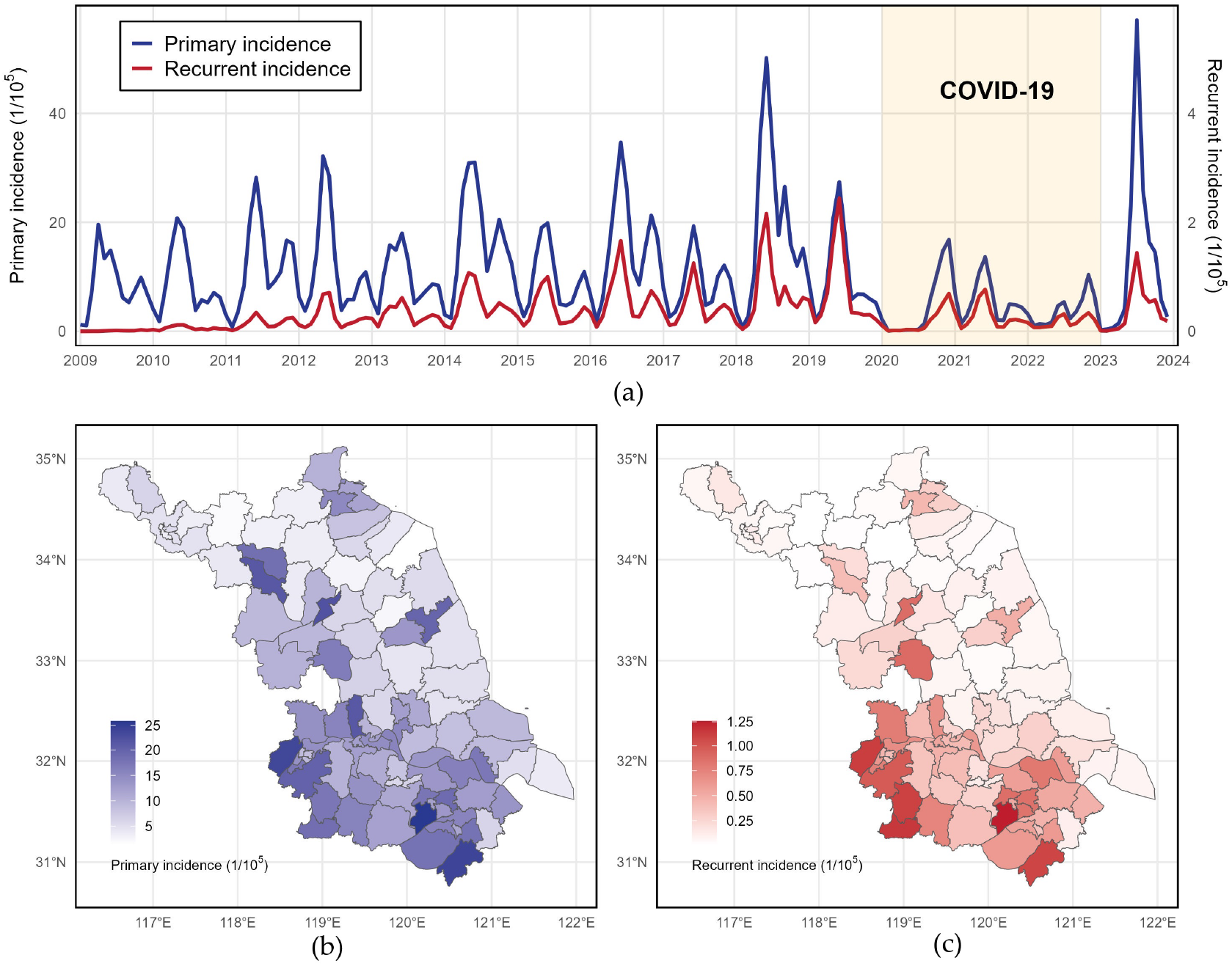
Spatiotemporal distribution of HFMD primary and recurrent incidence in Jiangsu, China during 2009–2023 (a). Cumulative incidence of primary and recurrent infection rate. Spatial distribution of primary incidence (b) and recurrent incidence (c) for the 14-year average.

We conducted a *χ*^2^ independence test to examine the association between specific demographic characteristics and infection type, revealing significant links for gender, age group, type of accommodation, and illness severity (Table 2). Among primary infection cases, the majority are males (58.0%) and children aged 1–2 years (25.3%). In contrast, reinfection cases are more prevalent among individuals aged 5+ years (1.0%). Cumulative incidence curves (Figure 3) illustrate the probability of recurrent infections for individuals in different sub-groups. Most reinfections occur within 2000 days of the initial infection, with males exhibiting a slightly higher probability of reinfection. Patients are grouped into six age categories (0–4 and 5+ years), and those first infected between 1–2 years old are most likely to have recurrent infections. Notably, patients with different severity levels of their first infection show different probabilities of recurrent infection. Critically infected patients have a reinfection probability of 0.06, significantly higher than the 0.04 observed in non-critically infected patients, and they also face a prolonged period of heightened reinfection risk.

**Table 2:** Demographic characteristics and *χ*^2^ independence test results for primary and recurrent HFMD infections.

| Characteristics | Primary infection | Reinfection | Total cases | *p*-value |
| --- | --- | --- | --- | --- |
| Gender |  |  |  | < 0.001 |
| Male | 885,354 (0.580) | 30,744 (0.020) | 916,098 (0.600) |  |
| Female | 591,940 (0.388) | 17,610 (0.012) | 609,550 (0.400) |  |
| Age group |  |  |  | < 0.001 |
| [0, 1) | 141,339 (0.093) | 114 (0.000) | 141,453 (0.093) |  |
| [1, 2) | 385,803 (0.253) | 3,726 (0.002) | 389,529 (0.255) |  |
| [2, 3) | 268,443 (0.176) | 7,904 (0.005) | 276,347 (0.181) |  |
| [3, 4) | 269,928 (0.177) | 11,540 (0.008) | 281,468 (0.184) |  |
| [4, 5) | 195,426 (0.128) | 10,170 (0.007) | 205,596 (0.135) |  |
| $\geq 5$ | 216,355 (0.142) | 14,900 (0.010) | 231,235 (0.152) | |
| Type |  |  |  | < 0.001 |
| Scattered | 895,921 (0.587) | 16,249 (0.011) | 912,170 (0.598) |  |
| Kindergarten | 503,041 (0.330) | 26,490 (0.017) | 529,531 (0.347) |  |
| School | 77,725 (0.051) | 5,611 (0.004) | 83,336 (0.055) |  |
| Unknown | 607 (0.000) | 4 (0.000) | 611 (0.000) |  |
| Critically ill |  |  |  | < 0.001 |
| 1 | 9,168 (0.006) | 178 (0.000) | 9,346 (0.006) |  |
| 0 | 1,467,652 (0.962) | 48,162 (0.032) | 1,515,814 (0.994) |  |
| Unknown | 474 (0.000) | 14 (0.000) | 488 (0.000) |  |
| Total | 1,477,294 (0.968) | 48,354 (0.032) | 1,525,648 (1.000) |  |

**Figure 3:**
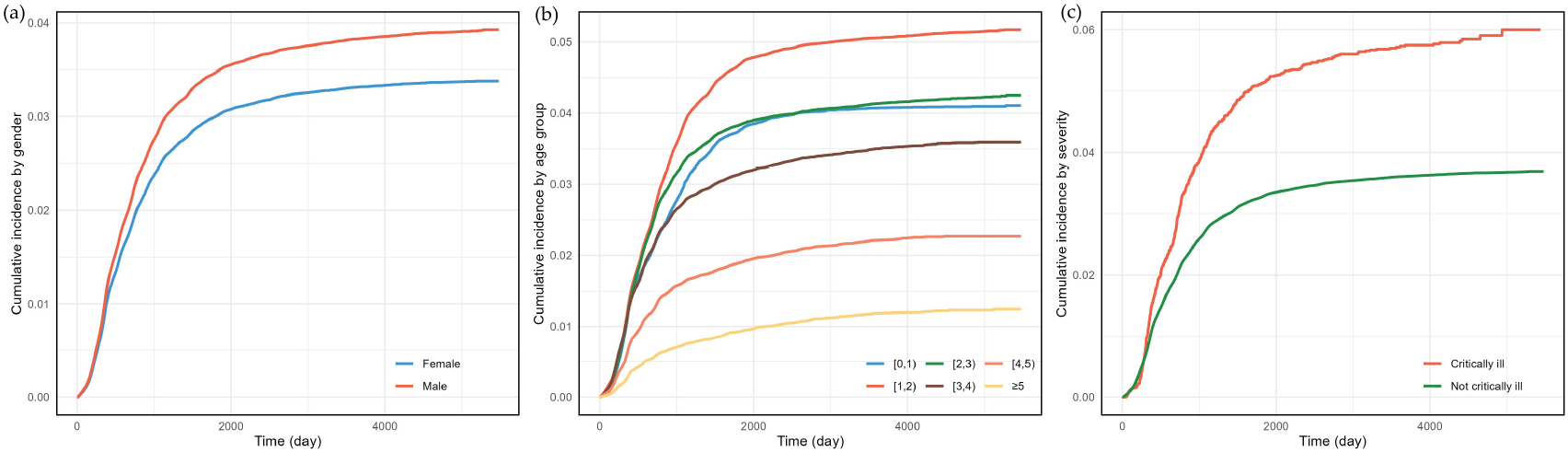
Cumulative incidence curves for different groups of gender, age, and severity of first infection.

### 4.2 Model results

Before establishing the Bayesian joint model, we tested multicollinearity using the Variance Inflation Factor (VIF), with all values below 2 (i.e., <10), indicating no multicollinearity (Salmeron et al., 2018). Model fitness was evaluated using the deviance information criterion (DIC), Watanabe-Akaike information criterion (WAIC), and the summative form of the logarithm score of conditional predictive ordinates (LS). Model 4, described in Section 3.1, is preferred since it has the lowest score for all three standards (DIC = 217,208, WAIC = 217,200, LS = 108,602) and is selected for further analysis. The calculation methods for the three standards and the fitness overview of other models are detailed in Appendix A.3. Based on the optimal model, the posterior mean of fixed effects for both sub-models and the 95% credible interval (CI) are summarized in Table 3. We identified significant factors influencing both primary and recurrent HFMD infections, including PM_2.5_, O_3_, NO_2_, wind speed, relative humidity, solar radiation, and COVID-19 restrictions.

**Table 3:** Posterior mean and 95% CI of fixed effects in the spatiotemporal joint model.

| Covariate | Primary infection model |  | Reinfection model |  |
| --- | --- | --- | --- | --- |
|  | Mean | 95% CI | Mean | 95% CI |
| PM<sub>2.5</sub> | −0.056 | (−0.085, −0.028) | −0.062 | (−0.119, −0.005) |
| O<sub>3</sub> | −0.226 | (−0.260, −0.191) | −0.099 | (−0.162, −0.035) |
| NO<sub>2</sub> | 0.167 | (0.133, 0.201) | 0.172 | (0.118, 0.227) |
| Temperature ≤ 27°C | 0.002 | (−0.007, 0.011) | −0.029 | (−0.054, −0.013) |
| Temperature > 27°C | −0.035 | (−0.058, −0.013) | 0.038 | (0.001, 0.074) |
| Wind speed | 0.056 | (0.041, 0.072) | 0.089 | (0.063, 0.115) |
| Precipitation | −0.099 | (−0.119, −0.078) | −0.026 | (−0.057, 0.005) |
| Relative humidity | 0.086 | (0.060, 0.113) | 0.130 | (0.084, 0.176) |
| Solar radiation | 0.067 | (0.027, 0.106) | 0.069 | (0.003, 0.136) |
| COVID-19 | −0.448 | (−0.562, −0.334) | −0.897 | (−1.523, −0.273) |
| Critically ill |  | – | 0.078 | (0.060, 0.096) |
| Scattered status | 0.932 | (0.911, 0.954) |  | – |

PM_2.5_ and O_3_ both negatively influence primary and recurrent infection. With one unit standard deviation (22.98 µg/m^3^) increase in PM_2.5_ concentration, primary infection incidence decreases by a factor of 0.95 (95% CI: 0.919, 0.972) and reinfection decreases by a factor of 0.94 (95% CI: 0.888, 0.995). Similarly, with one unit standard deviation (31 µg/m^3^) increase in O_3_ concentration, incidence becomes 0.80 (95% CI: 0.771, 0.826) times and 0.91 (95% CI: 0.850, 0.966) times lower for primary and recurrent infection respectively. On the other hand, NO_2_ shows a positive association with HFMD incidence, with every 12.35 µg/m^3^ increase in NO_2_ concentration resulting in an 18% increase in primary infections (1.18, 95% CI: 1.142, 1.223) and a 19% increase in recurrent infections (1.19, 95% CI: 1.125, 1.254).

Among meteorological variables, we fitted temperature using a cut-point model (Bauer and Wakefield, 2018) with the cut-point at 27 °C to reflect its non-linear association with HFMD incidence (Huang et al., 2018a). Primary infection showed a positive association with temperature below 27 °C, such that a 1 °C increase in temperature was associated with a 1.002-fold (95% CI: 0.993, 1.011; not significant) increase in disease incidence. In contrast, the incidence decreased as the temperature rose above 27 °C (−0.035, 95% CI: −0.058, −0.013). The influence of temperature, however, followed a different trend in the reinfection sub-model, with a negative association below 27 °C (−0.029, 95% CI: −0.054, −0.013) and a positive association above 27 °C (0.038, 95% CI: 0.001, 0.074). Wind speed, relative humidity, and solar radiation positively correlated with primary and recurrent infection. Higher precipitation was significantly associated with lower primary infection incidence, with a 1.53 mm (one unit standard deviation) increase in precipitation associated with a 0.099-fold (95% CI: 0.078, 0.119) decrease in incidence. Precipitation also negatively impacted reinfection incidence but was not statistically significant (−0.026, 95% CI: −0.057, 0.009).

In the reinfection sub-model, the severity of the primary infection is suggested to impact recurrent incidence. One unit standard deviation (0.25%) increase in the proportion of critically infected patients for their first infection tends to raise reinfection incidence by 8% (95% CI: 6%, 10%). In the primary infection model, individuals categorized as scattered are identified as having a higher risk of infection. For every 10.3% increase in the proportion of scattered patients, primary infection incidence is expected to rise by a factor of 2.54 (95% CI: 2.49, 2.60). During the COVID-19 period, both primary and recurrent infections show significant reductions, with the incidence decreasing to 0.639 (95% CI: 0.570, 0.716) and 0.408 (95% CI: 0.218, 0.761) times, respectively, compared to a non-COVID period.

Table 4 displays posterior estimates of model hyperparameters, including precision for spatial variation *τ* ^*s*^ in the BYM2 model, precision for monthly and yearly variation *τ* ^*m*^ and *τ* ^*y*^ in the RW1 model, and the precision for spatiotemporal interaction in two sub-models 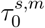 and 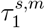.Sharing coefficients for the monthly, yearly, and spatial latent effects 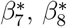 and 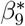 are also shown. The monthly random effect accounts for the most variability with a posterior mean precision of 9.434 (variance = 1/9.434). The spatiotemporal interaction in the primary infection model has stronger variability than in the reinfection model, with posterior means 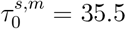 and 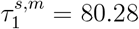, respectively. The yearly random effect shows a smaller precision of *τ* ^*y*^ = 195.17, and the spatial component accounts for the least variability with a variance of 1/2396.75. All three sharing coefficients are significantly greater than 0. The most significant sharing coefficient, 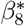 (mean = 5.774), indicates a substantial similarity in the yearly trends between primary infection incidence and reinfection incidence, while the monthly variation sharing coefficient, 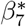 (mean = 1.823), suggests a weaker but still significant similarity. Additionally, both sub-models exhibit similarities in spatial patterns, with a mean 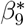 of 0.99.

**Table 4:** Posterior estimates of model hyperparameters in the spatiotemporal joint model.

| Parameter | Mean | SD | 2.5% quantile | 50% quantile | 97.5% quantile |
| --- | --- | --- | --- | --- | --- |
| $\tau^s$ (BYM2) | 2396.75 | 5430.00 | 0.43 | 56.92 | 1220.00 |
| $\tau^m$ (RW1) | 9.43 | 4.13 | 3.85 | 8.62 | 19.80 |
| $\tau^y$ (RW1) | 195.17 | 76.50 | 85.90 | 181.87 | 382.00 |
| $\tau_0^{s,m}$ (Interaction) | 35.50 | 4.22 | 28.29 | 35.14 | 44.90 |
| $\tau_1^{s,m}$ (Interaction) | 80.28 | 24.80 | 42.06 | 76.81 | 139.00 |
| $\beta_7^*$ (Shared monthly) | 1.82 | 0.08 | 1.68 | 1.82 | 1.97 |
| $\beta_8^*$ (Shared yearly) | 5.77 | 0.18 | 5.43 | 5.77 | 6.14 |
| $\beta_9^*$ (Shared spatial) | 0.99 | 0.32 | 0.33 | 1.00 | 1.59 |

### 4.3 Non-linear results

The estimated non-linear effects of air pollution and meteorological factors on HFMD incidence are shown in Figure 4, where similar effects are observed for primary and recurrent infection. For PM_2.5_, the posterior mean is almost negative for concentration below 50 µg/m^3^, indicating a negative impact on primary and recurrent incidence, becomes positive between 50 and 70 µg/m^3^, and then gradually declines with fluctuations. A positive but weakening association is observed for O_3_ below 90 µg/m^3^, after which the association turns negative and strengthens for primary infection while remaining relatively stable for reinfection. In most cases, NO_2_ concentration is positively associated with primary and recurrent incidence, with a stronger negative influence on primary incidence for concentration greater than 80 µg/m^3^.

**Figure 4:**
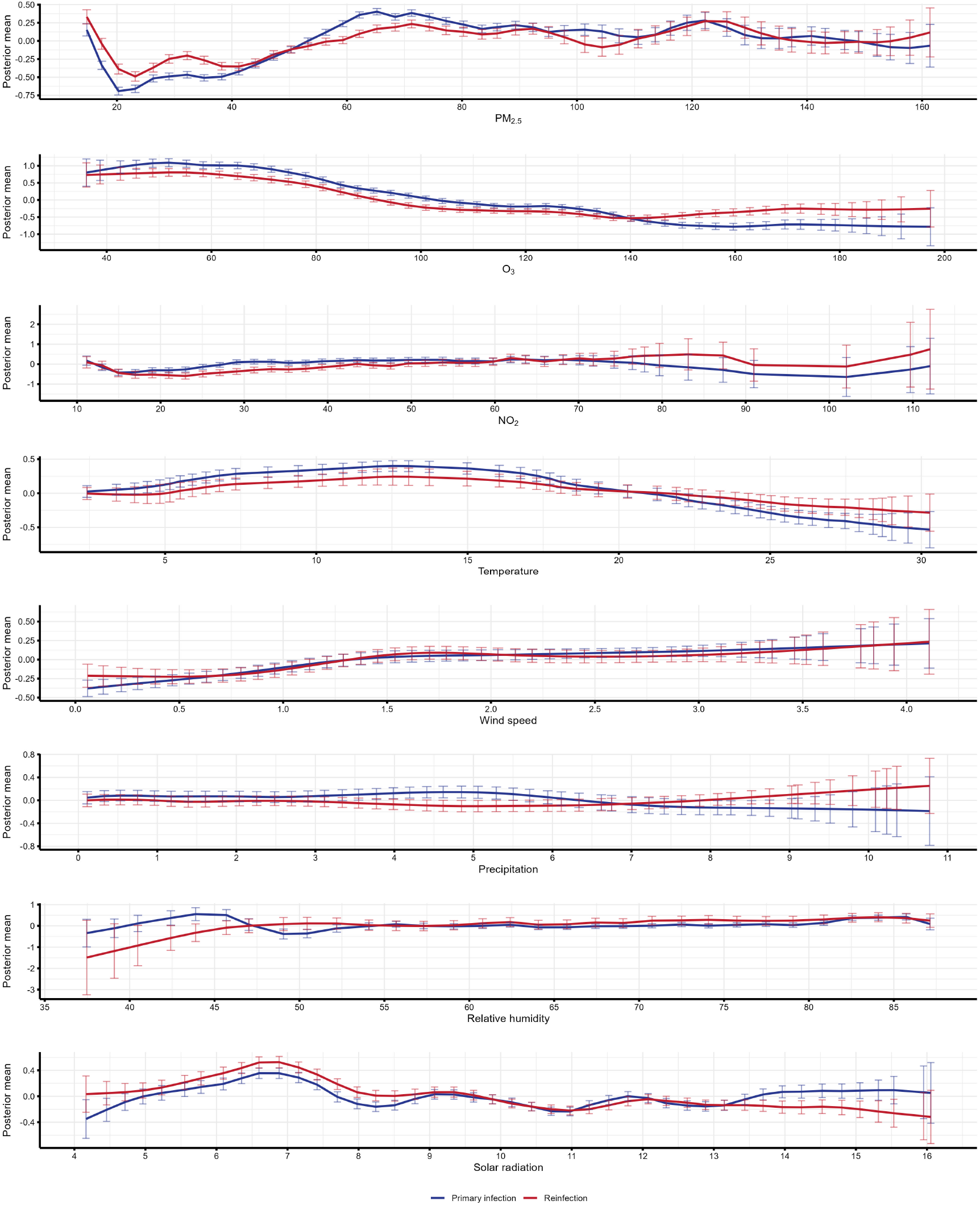
Estimated non-linear effect of air pollution and meteorological factors on primary and recurrent incidence.

The posterior mean for temperature is positive below 20 °, and relative humidity is positively associated with HFMD incidence when it exceeds 55%, with primary infection showing more significant fluctuation than reinfection. For wind speed, a similar pattern is observed around 1.25 m/s, speeds below this threshold are negatively correlated with HFMD incidence, while those above it show a positive correlation. Non-linear effects of precipitation demonstrate slightly different trends for primary and recurrent incidence, such that below 7 mm, it is positive for primary infection but negative for reinfection, and above 7 mm, the effect reverses. Finally, the impact of solar radiation experiences a series of fluctuations, with the greatest positive effect on both primary and recurrent infections observed between 6 MJ/m^3^ and 7 MJ/m^3^.

### 4.4 Evaluation of joint modelling and sensitivity analysis

We assessed estimation accuracy for joint and independent models. Table 5 displays the standard deviation for ten common fixed effects in independent and joint models. In the primary infection model, the standard deviation of all covariates decreased in the joint model, ranging from 0.004 (for O_3_) to 0.107 (for COVID-19). In contrast, the improvements in standard deviation were smaller in the reinfection model, likely due to the reduced data size, with the largest difference of 0.051 for COVID-19 (a reduction of 13.5%). These results indicate more accurate and unbiased coefficient estimation for the joint model, confirming the joint modeling technique’s appropriateness.

**Table 5:** Standard deviation of fixed effects in joint model and independent model.

| Covariate | Independent model |  | Joint model |  |
| --- | --- | --- | --- | --- |
|  | Primary | Recurrent | Primary | Recurrent |
| PM<sub>2.5</sub> | 0.019 | 0.029 | 0.015 | 0.029 |
| O<sub>3</sub> | 0.023 | 0.033 | 0.018 | 0.032 |
| NO<sub>2</sub> | 0.019 | 0.028 | 0.017 | 0.028 |
| Temperature$\leq 27^{\circ}\text{C}$ | 0.006 | 0.010 | 0.005 | 0.008 |
| Temperature$> 27^{\circ}\text{C}$ | 0.013 | 0.021 | 0.011 | 0.019 |
| Precipitation | 0.011 | 0.016 | 0.010 | 0.016 |
| Relative humidity | 0.015 | 0.024 | 0.014 | 0.023 |
| Wind speed | 0.009 | 0.013 | 0.008 | 0.013 |
| Solar radiation | 0.024 | 0.037 | 0.020 | 0.034 |
| COVID-19 | 0.167 | 0.378 | 0.060 | 0.327 |

The sensitivity analysis of priors shows that the results are consistent in both direction and significance (Tables A.2 and A.3).

## 5 Discussion

This study innovatively applied a joint model to simultaneously analyze primary and recurrent HFMD infections, capturing their shared and unique spatiotemporal patterns. By employing the joint model, we identified similarities in the spatial distribution and temporal trends (monthly and yearly) of both infection types, with the most significant variation occurring at the yearly level. This aligns with the idea that smaller time units exhibit more significant variability and flexibility, reducing the likelihood of two infection patterns matching. Utilizing detailed county-level data from 95 counties in Jiangsu province, China, we conducted a granular analysis of HFMD incidence, allowing for greater spatial variability to be detected and reflected, resulting in smaller sharing coefficients. To validate the joint model, we compared the standard deviations of the fixed effects between the joint and independent models. Our findings were consistent with Agasa et al. (2024), demonstrating that one strength of joint modeling on infectious disease is reduced uncertainty and estimation bias, particularly when there is a possibility of under-reporting. The joint model captures the association between recurrent and primary infections, reducing uncertainty in both sub-models and improving estimation accuracy.

All three air pollution variables exhibit significant linear association with HFMD incidence in two sub-models. PM_2.5_ and O_3_ are identified as suppressive factors of HFMD transmission, which is also reflected in the research from Zhang et al. (2022) that higher incidence occurs with better air quality. While some existing literature suggests that higher concentrations of air pollution particles like PM_2.5_ can facilitate viral transmission in the air, leading to increased disease incidence (Marquès and Domingo, 2022), the non-linear association of PM_2.5_ reported by Qian et al. (2023) and Peng et al. (2022) and O_3_ reported by Luo et al. (2024) and Yan et al. (2019) show relative risk less than 1 in most cases, which corresponds to the negative linear association and non-linear trend displayed in our study. As PM_2.5_ and O_3_ concentrations rise, people are more likely to become aware and take action to protect themselves from pollutants, such as wearing masks or reducing outdoor activities. While these actions primarily reduce exposure to harmful pollutants, they may indirectly help reduce the overall burden on the immune system, potentially lowering susceptibility to infections like HFMD (Wu et al., 2022). In contrast, NO_2_ is positively related to HFMD incidence, consistent with the research by Gu et al. (2020). Studies from Peng et al. (2022) and Luo et al. (2024) demonstrate the non-linear trend of the influence of NO_2_ with relative risk geater than 1 in most cases, aligning with our result. Higher concentrations of NO_2_ can damage children’s respiratory systems by causing airway inflammation, impairing lung function, and weakening immune responses, which may increase their susceptibility to HFMD (Dondi et al., 2023).

Regarding average temperature, we observed opposite trends for primary and recurrent infections, with a cut-off point at 27 °. Specifically, rising temperatures are generally associated with higher primary incidence until reaching 27°C, after which higher temperatures become less conducive to virus survival and transmission (Huang et al., 2018a). For recurrent infections, the effect is negative below 27 ° and positive above 27 °, aligning with the findings from related studies in Guangdong province from Yan et al. (2019) and Chen et al. (2020). Such distinctions may be attributed to different types of virus-infected in primary and recurrent cases and their responses to the temperature change (Dong et al., 2016). A study from Huang et al. (2018b) shows different probabilities of reinfection caused by different types of viruses, with primary infection caused by EV-A71 and CV-A16. Additionally, Qi et al. (2020) indicated in their research that much higher risk was related to EV-A71 on cold days than other virus serotypes. Further research may be conducted to identify specific effects of temperature or other meteorological factors on different serotypes.

Meanwhile, non-linear analysis suggests that temperature has a more substantial effect on primary incidence than on recurrent infections, though these differences require further verification in future studies. Wind speed, relative humidity, and solar radiation are suggested to influence both primary and recurrent infections positively. To some extent, higher wind speed may boost virus transmission and increase the risk of infection (Song et al., 2018). Similarly, higher relative humidity increases virus accumulation and may restrict body metabolism, making children more susceptible to HFMD (Luo et al., 2020). The effect of solar radiation on HFMD has not been fully understood, but our findings correspond with Wang et al. (2016), suggesting that exposure to ultraviolet radiation may damage the immune system and weaken resistance to infections (Norval, 2001). On the other hand, precipitation is negatively associated with HFMD incidence, as higher rainfall may reduce virus transmission by decreasing airborne viral concentration, limiting outdoor activities, and promoting environmental cleaning (Guo et al., 2023).

Our findings suggest that recurrent infections are significantly associated with the severity of the first infection, as severe initial infections may cause cardiac dysfunction and neurological damage (Ji et al., 2024), increasing the likelihood of reinfection. Additionally, lifestyle factors, such as frequent exposure to crowded environments or poor hygiene practices (Li et al., 2021), may further elevate the risk of reinfection, especially in individuals with a compromised immune system. A higher incidence of primary infection is found among scattered children as indicated by Hu et al. (2024). This is probably due to the younger age group of scattered children, as HFMD most commonly affects young children whose immune systems are still developing and thus less capable of effectively combating the virus (U.S. Centers for Disease Control and Prevention, 2024). Finally, the incidence of both types of infections is significantly low during COVID-19. On the one hand, COVID-19 restrictions, including national lockdowns and quarantine policies, may reduce the probability of contacting a transmission source (Zhao et al., 2022). On the other hand, The government’s focus on COVID-19 control likely diverted resources from monitoring HFMD, leading to under-reporting (Li et al., 2024). Factors include delayed detection, reduced public attention, parents avoiding healthcare due to COVID-19 fears, and changes in reporting systems prioritizing COVID-19 cases.

This study has several limitations. First, while it primarily focuses on the environmental impact on HFMD incidence, factors such as breastfeeding status (Lin et al., 2014), vaccination (Du et al., 2021), and virus type (Xu et al., 2015)—which may influence susceptibility— were not analyzed due to data constraints. Second, this study explores risk factors and associations between primary and recurrent infections at the population level, but it cannot capture potential biases arising from individual-level heterogeneity, future research should focus on more detailed individual-level analyses to provide deeper insights.

## Data Availability

All data produced in the present study are available upon reasonable request to the authors and with permission of the Jiangsu Provincial Center for Disease Control and Prevention.

## A Appendix

### A.1 BYM2 model

The Besag York Mollié (BYM) model is a useful technique for disease mapping in that it combines different types of spatial random effects. The conditional auto-regressive model proposed by Besag et al. (1991) suggests that the spatial effect of a particular region depends on its neighboring area, where a spatial auto-correlation is included that assumes nearby locations have more similarities than locations that are farther apart. Recall that an intrinsic conditional auto-regressive (ICAR) model is a normal distributed random field {*u*_*i*_, *i* ∈ *I*} with a mean equal to the average of its neighbors, and variance decreases as the number of neighbors, denoted *d*_*i*_, increases (Rue et al., 2009). Namely,

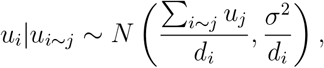

where *σ*^2^ is an unknown variance and *i* ∼ *j* stands for the neighbour relationship for *i* ≠ *j*. The proposed BYM model consists of the structured spatial effect illustrated in the aforementioned ICAR model and an unstructured component representing independent region-specific variations.

Compared with the BYM model, the BYM2 model was proposed by Simpson et al. (2017) with a clearer interpretation of the new parameters, allowing for the assignment of sensible hyperparameters to each. BYM2 model introduced a mixing parameter *ϕ* to combine the structured random effect *u* and unstructured random effect *v* as

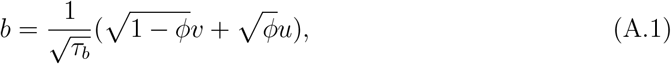

where *τ*_*b*_ stands for the precision, and the mixing parameter *ϕ* (0 ≤ *ϕ* ≤ 1) controls the proportion of *u* and *v* in the random effects, which is as well the extent the structured component and unstructured component contributed respectively. The parameter *τ*_*b*_ *>* 0 controls the contribution amount of spatial random effect *b*. As a result, the random effect consists of the purely structured component when *ϕ* = 1 and an unstructured component when *ϕ* = 0. The penalized complexity (PC) prior is applied to penalize model complexity via formulating prior distributions of parameters *τ* and *ϕ*. The base model assumes constant relative risk (corresponding to *τ*_*b*_ close to zero) across the whole area, while the opposite level refers to completely varying risk.

### A.2 RW1 model

As one of the latent effects available in R-INLA, first-order Gaussian random walks (RW1) help capture nonlinear covariate effects or temporal correlation (Gómez-Rubio, 2020). Given a vector of Gaussian observations **u** = (*u*_1_, …, *u*_*n*_), the RW1 is defined assuming that increments Δ*u*_*i*_ = *u*_*i*_ − *u*_*i*−1_ follow a Gaussian distribution with zero mean and precision *τ*

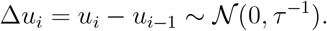

The current observation is assumed to be previous observations with a random noise. The density for **u** is derived from its *n* − 1 increments as

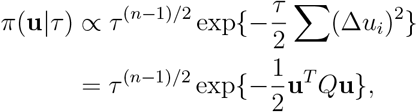

where *Q* = *τ R* and *R* are the structure matrix reflecting the neighborhood structure of the model. The RW1 model enables a cyclic setting in which the last node *u*_*n*_ is the neighbor of *u*_*n*−1_ and *u*_1_.

### A.3 Evaluation criteria

Deviance information criterion (DIC), proposed by Spiegelhalter et al. (2002), is a widely applied criterion for model choice defined as 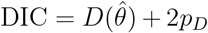,where 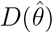 is the deviance function with estimate *θ*, and 2*p*_*D*_ is the effective number of parameters. Watanabe-Akaike information criterion (WAIC) (Watanabe, 2013), also known as widely applicable information criterion, is the generalized version of the Akaike information criterion (AIC) onto singular statistical models. It is defined as

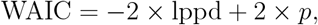

where lppd is the log point-wise predictive density, and *p* is the effective number of parameters. WAIC is more desirable for averaging over the posterior distribution rather than conditioning on a point estimate. The conditional predictive ordinates (CPO) (Pettit, 1990) is the leave-one-out cross-validation predictive density *p*(*y*_*i*_|*y*_*i*−1_) = ∫ *p*(*y*_*i*_|*θ*)*p*(*θ*|*y*_−*i*_)*dθ*, that estimates probability of observing *y*_*i*_ given the observed *y*_−*i*_ for each *i*. CPO requires only a sample from the posterior distribution without additional simulation and is computationally efficient. In this study, we use CPO with its summative form of logarithm score LS (Zhang et al., 2023)

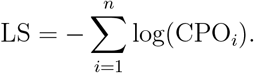

**Table A1:** Model performance.

| Model | DIC | WAIC | LS |
| --- | --- | --- | --- |
| Model 1 | 225,099 | 225,105 | 112,552 |
| Model 2 | 224,071 | 224,082 | 112,041 |
| Model 3 | 218,077 | 218,094 | 109,047 |
| **Model 4** | **217,208** | **217,200** | **108,602** |

### A.4 Sensitivity analysis results

**Table A2:** Posterior mean and 95% CI of fixed effects under prior settings in temporal effect in the spatiotemporal joint model.

| Covariate | Primary infection model |  | Reinfection model |  |
| --- | --- | --- | --- | --- |
|  | Mean | 95% CI | Mean | 95% CI |
| PM<sub>2.5</sub> | −0.056 | (−0.085, −0.027) | −0.062 | (−0.119, −0.005) |
| O<sub>3</sub> | −0.226 | (−0.261, −0.191) | −0.099 | (−0.162, −0.035) |
| NO<sub>2</sub> | 0.167 | (0.133, 0.201) | 0.173 | (0.118, 0.227) |
| Temperature≤ 27°C | 0.002 | (−0.007, 0.011) | −0.029 | (−0.045, −0.014) |
| Temperature> 27°C | −0.035 | (−0.057, −0.013) | 0.038 | (−0.001, 0.075) |
| Wind speed | 0.056 | (0.041, 0.072) | 0.089 | (0.063, 0.115) |
| Precipitation | −0.099 | (−0.119, −0.079) | −0.026 | (−0.057, 0.005) |
| Relative humidity | 0.087 | (0.060, 0.113 ) | 0.130 | (0.084, 0.176) |
| Solar radiation | 0.067 | (0.027, 0.106) | 0.070 | (0.003, 0.136) |
| COVID-19 | −0.448 | (−0.574, −0.323) | −0.898 | (−1.591, −0.207) |
| Critically ill |  | — | 0.078 | (0.060, 0.096) |
| Scattered status | 0.932 | (0.911, 0.954) |  | — |

**Table A3:** Posterior mean and 95% CI of fixed effects under prior settings in spatial effect in the spatiotemporal joint model.

| Covariate | Primary infection model |  | Reinfection model |  |
| --- | --- | --- | --- | --- |
|  | Mean | 95% CI | Mean | 95% CI |
| PM<sub>2.5</sub> | −0.057 | (−0.085, −0.028) | −0.061 | (−0.118, −0.005) |
| O<sub>3</sub> | −0.225 | (−0.260, −0.190) | −0.099 | (−0.163, −0.036) |
| NO<sub>2</sub> | 0.167 | (0.133, 0.201) | 0.172 | (0.117, 0.226) |
| Temperature≤ 27°C | 0.002 | (−0.007, 0.011) | −0.029 | (−0.045, −0.013) |
| Temperature> 27°C | −0.035 | (−0.057, −0.013) | 0.038 | (0.001, 0.074) |
| Wind speed | 0.056 | (0.041, 0.072) | 0.089 | (0.063, 0.115) |
| Precipitation | −0.099 | (−0.119, −0.078) | −0.026 | (−0.057, 0.005) |
| Relative humidity | 0.087 | (0.060, 0.113) | 0.129 | (0.084, 0.175) |
| Solar radiation | 0.066 | (0.027, 0.105) | 0.070 | (0.003, 0.136) |
| COVID-19 | −0.448 | (−0.563, −0.333) | −0.897 | (−1.534, −0.262) |
| Critically ill |  | — | 0.078 | (0.060, 0.096) |
| Scattered status | 0.932 | (0.911, 0.954) |  | — |

## Declaration

### Ethics approval and consent to participate

The study is approved by the Xi’an Jiaotong-Liverpool University Research Ethics Committee (0010000089620241213145739).

### Consent for publication

Not applicable.

### Availability of data and materials

The data that support the findings of this study are available from the Jiangsu Provincial Center for Disease Control and Prevention, but restrictions apply to the availability of these data, which were used under license for the current study and so are not publicly available.

Data are, however, available from Liguo Zhu upon reasonable request and with permission of the Jiangsu Provincial Center for Disease Control and Prevention.

### Authors’ contributions

Wang Y., Bao C. and Ling C. contributed to the research design, conceptualization and validation; Zhu L. contributed to resource and validation; Wang W. contributed to writing (original draft), formal analysis and visualization; Tang Y. contributed to writing (review & editing), methodology and visualization; Ji H. contributed to methodology, resource and validation; Zhu H. contributed to data curation, resource and validation; Liu W. and Wang K. contributed to methodology and validation. All authors participated in the revision and approved the manuscript.

## Acknowledgements

We thank the editors and anonymous reviewers for their helpful remarks.

